# Sex Differences in the Independent and Combined Effects of Genomic and Exposomic Risks for Schizophrenia on Distressing Psychotic Experiences: Insights from the ABCD Study

**DOI:** 10.1101/2025.05.09.25327350

**Authors:** Thanavadee Prachason, Angelo Arias-Magnasco, Bochao Danae Lin, Jim van Os, Bart P.F. Rutten, Lotta-Katrin Pries, Sinan Guloksuz

**Author notes:** Corresponding author: Sinan Guloksuz, Department of Psychiatry and Neuropsychology, School for Mental Health and Neuroscience, Maastricht University Medical Center, P.O. Box 616 6200 MD Maastricht, The Netherlands. Shared last author (Lotta-Katrin Pries, Sinan Guloksuz).

## Abstract

**Purpose:** To investigate sex-dependent effects of polygenic risk (PRS-SCZ) and exposome score (ES-SCZ) for schizophrenia, both independently and jointly, on distressing psychotic experiences (PEs) in early adolescents.

**Method:** Baseline to 3-year follow-up data of the Adolescent Brain and Cognitive Development Study (ABCD) were used. PRS-SCZ and ES-SCZ were calculated to assess cumulative genetic and environmental (childhood adversity, cannabis use, hearing impairment, and winter births) risk for schizophrenia, respectively. The primary outcome was past-month distressing PEs at the 3-year follow-up. Secondary outcomes included distressing PEs across four yearly assessments: lifetime (≥1 wave), repeated (≥2 or ≥3 waves), and persisting (≥4 waves). Sex-stratified multilevel logistic regression models were used to test the independent and joint associations of binary modes (>75th percentile) of PRS-SCZ (PRS-SCZ_75_) and ES-SCZ (ES-SCZ_75_) on the outcomes. As sensitivity analysis, the sex-stratified analyses were repeated on a randomly selected unrelated sample, and the coefficients of males and females were compared.

**Results:** PRS-SCZ_75_ was not associated with past-month distressing PEs in either sex but significantly associated with lifetime and repeated (≥2 waves) distressing PEs only in females. However, a significant sex difference was only detected for lifetime distressing PEs. ES-SCZ_75_ was significantly associated with past-month distressing PEs and all secondary outcomes without significant sex differences. No additive interaction between ES-SCZ_75_ and PRS-SCZ_75_ was detected for any PE outcomes.

**Conclusion:** The influence of genomic risk for schizophrenia on distressing PEs might be sex-dependent, whereas that of the exposomic risk was universal in early adolescence. Further studies in larger samples are needed.

**Article Highlights:**

▢ Genomic risks for schizophrenia were associated with distressing PEs only in female adolescents.
▢ Environmental risks for schizophrenia were associated with distressing PEs regardless of sex in early adolescence.
▢ Dose-response effects of the environmental risks on PE persistence support them as potential preventive targets.
▢ A sex-informed approach is needed to better understand genetic and environmental contributions to psychosis expression.

## Introduction

Psychotic experiences (PEs), including phenomena such as hallucinations and delusions, often emerge during childhood and adolescence(Healy et al. 2019). Early PEs do not only increase the risk of later psychotic spectrum disorders (PSD), but are also associated with other severe mental health outcomes in adulthood, particularly when persisting over an extended period(Kelleher et al. 2012; Staines et al. 2023). Therefore, understanding the factors contributing to PEs in youth is essential to optimize early intervention strategies.

The development of psychosis is influenced by gene-environment interaction(Wahbeh & Avramopoulos 2021; Zwicker et al. 2018). To quantify polygenetic risk, the polygenic risk score for schizophrenia (PRS-SCZ) has been developed as a weighted sum of schizophrenia risk alleles(International Schizophrenia Consortium et al. 2009) that could predict PSD and other mental health outcomes in both clinical and general populations(Mistry et al. 2018; Pries et al. 2020). Much like genetic risks, environmental exposures have been shown to influence mental health through complex and cumulative effects(Guloksuz et al. 2018a; Guloksuz et al. 2018b). The exposome score for schizophrenia (ES-SCZ)(Pries et al. 2021b; Pries et al. 2019), a weighted aggregated score of important environmental risk factors for schizophrenia (i.e., childhood adversity, cannabis use, hearing impairment, winter birth, and bullying), has been developed and shown to predict psychosis expression from subtle to severe phenotypes(Pries et al. 2021a; Pries et al. 2020). Furthermore, research suggests that ES-SCZ and PRS-SCZ jointly influence PSD(Pries et al. 2020) and PEs in early adolescence(Di Vincenzo et al. 2025), highlighting the intricate interplay between genetic and environmental factors in shaping psychosis risk.

Adding further complexity to the pathoetiology of psychosis, emerging evidence highlights the pivotal role of sex in shaping the development, progression, and outcomes of PSD(Morgan et al. 2008). While prior studies suggest sex differences in how individual environmental factors affect psychosis expression(Pence et al. 2022), research on sex-specific effects of exposomic risks remains limited. Notably, ES-SCZ has shown sex-specific associations with physical health, with worse outcomes observed in women(Paquin et al. 2023). However, its role for PEs during adolescence remains unexplored. Likewise, research on the sex-specific effects of PRS-SCZ especially on PEs is scarce(Docherty et al. 2020; Mas-Bermejo et al. 2023), creating a crucial gap in our understanding of adolescent mental health.

In this study, sex differences in the independent and additive effects of PRS-SCZ and ES-SCZ on distressing PEs were investigated in a young general population cohort.

## Methods

### Participants

The ABCD Study is an ongoing multisite cohort study conducted in the United States, following the development of 11,876 children from early adolescence to young adulthood(Barch et al. 2018). The current study utilized data from baseline to 3-year follow-up of the ABCD cohort, Data Release 5.1 (see Acknowledgements). Inclusion criteria for the current analyses were of European descent, having good-quality genotyping data, and having a binary sex reported at birth. Samples with incomplete data on any variable were excluded from related analyses, resulting in 2,401 females and 2,721 males for the main analysis. Missing reports are shown in Online Resource 1. The ABCD study was approved by the Centralized Institutional Review Board (IRB) of the University of California-San Diego and local research site IRBs in accordance with their IRB-approved protocols, state regulations, and local resources. Written informed consent and assent were derived from participating parents/caregivers and adolescents, respectively(Barch et al. 2018).

### Measurements Psychotic experiences

PEs were self-reported by adolescents using the 21-item Prodromal Questionnaire-Brief Child Version. This scale assesses PEs such as unusual thought content and perceptual abnormality over the past month(Loewy et al. 2011). Following previous research(Di Vincenzo et al. 2025; Karcher et al. 2022), ‘distressing PEs’ were defined as ≥1 PEs with a distressing score ≥3 out of a 5-point scale. The primary outcome was distressing PEs reported at 3-year follow-up (hereafter ‘past-month distressing PEs’). As secondary outcomes, four variables indicating the presence of distressing PEs from baseline to 3-year follow-up were generated at varying thresholds of persistence: distressing PEs present in ≥1 waves (hereafter ‘lifetime distressing PEs’), ≥2 waves (hereafter ‘repeating distressing PEs ≥2 waves’), ≥3 waves (hereafter ‘repeating distressing PEs ≥3 waves’), and all 4 waves (hereafter ‘persisting distressing PEs’).

### Exposome score for schizophrenia

ES-SCZ was calculated in alignment with previous reports(Di Vincenzo et al. 2025; Pries et al. 2019). Nine environmental factors, comprising childhood adversity (emotional and physical neglect; emotional, physical, and sexual abuse), bullying, cannabis use, winter birth, and hearing impairment, were taken from baseline to the 2-year follow-up assessments of the cohort. Each environmental factor was dichotomized to indicate lifetime exposure. Detailed information on how each environmental factor was derived was described in our previous report(Di Vincenzo et al. 2025) and Online Resource 1. Following previous research(Pries et al. 2019), ES-SCZ was calculated by adding the nine exposures multiplied by their weighted risks for schizophrenia.

### Polygenic risk score for schizophrenia

PRS-SCZ was generated for participants who passed the genetic and sample quality control. Detailed descriptions were provided in Online Resource 1. Briefly, PRS-SCZ was constructed using a Bayesian framework method that utilizes continuous shrinkage (cs) on SNP effect sizes(Ge et al. 2019) derived from the most recent schizophrenia GWAS (European subsample) based on 53,386 cases and 77,258 controls(Trubetskoy et al. 2022). The 1000 Genomes Project European Sample (https://github.com/getian107/PRScs) was used as an external linkage disequilibrium reference panel. To compute posterior effect sizes, the default settings of PRS-cs-auto were used (Online Resource 1). PRS-SCZ was then constructed based on 742,011 variants that pass quality control, using ‘--score’ function and the SUM modifier in PLINK1.9. Finally, PRS-SCZ was standardized before being used in further analyses.

### Statistical analysis

All analyses were conducted using Stata (version 16.1). Following prior research(Di Vincenzo et al. 2025), PRS-SCZ and ES-SCZ were dichotomized at the 75th percentile (hereafter PRS-SCZ_75_ and ES-SCZ_75_, respectively). Multilevel logistic regression models, accounting for study site and family structure, were used to examine the associations of PRS-SCZ_75_ and ES-SCZ_75_ with PEs. Additive interactions between PRS-SCZ_75_ and ES-SCZ_75_ were assessed using the relative excess risk due to interaction (RERI), calculated via the delta method using Stata’s “nlcom” command. A RERI >0 indicates an interaction effect beyond the sum of genomic and exposome risk states. Sex-stratified analyses were applied separately for the primary outcome (past-month distressing PEs) and each secondary outcome (lifetime, repeating ≥2 or 3 waves, and persisting distressing PEs). Model 1 was adjusted for age, and Model 2 was further adjusted for family income and parental education. Models including PRS-SCZ_75_ were adjusted for the first ten genetic principal components (Online Resource 1). As a sensitivity analysis, we randomly selected one participant per family to create an unrelated sample. Then, sex-stratified multilevel regression analyses were applied before the sex differences in the independent and joint associations of PRS-SCZ_75_ and ES-SCZ_75_ were determined using Chow’s test(Chow 1960).

## Results

Sample characteristics and the prevalence of distressing PEs are shown in Table 1.

**Table 1.** Participant characteristics.

| Characteristics at baseline | Males (N = 2,721) |  | Females (N = 2,401) |  |
| --- | --- | --- | --- | --- |
|  | Mean | SD | Mean | SD |
| Age (years) | 9.95 | 0.63 | 9.91 | 0.63 |
| Parental educational attainment (years) | 18.2 | 1.7 | 18.2 | 1.7 |
|  | **n** | **%** | **n** | **%** |
| Family income |  |  |  |  |
| < \$5,000 | 16 | 0.6 | 12 | 0.5 |
| \$5,000-\$11,999 | 24 | 0.9 | 10 | 0.4 |
| \$12,000-\$15,999 | 16 | 0.6 | 14 | 0.6 |
| \$16,000-\$24,999 | 37 | 1.4 | 41 | 1.8 |
| \$25,000-\$34,999 | 67 | 2.6 | 60 | 2.6 |
| \$35,000-\$49,999 | 143 | 5.5 | 125 | 5.4 |
| \$50,000-\$74,999 | 362 | 13.9 | 306 | 13.3 |
| \$75,000-\$99,999 | 423 | 16.3 | 418 | 18.1 |
| \$100,000-\$199,999 | 1,104 | 42.4 | 954 | 41.3 |
| ≥ \$200,000 | 410 | 15.8 | 370 | 16.0 |
| **Lifetime exposure up to 2-year follow-up** | **n** | **%** | **n** | **%** |
| Physical abuse | 25 | 0.9 | 13 | 0.5 |
| Emotional abuse | 24 | 0.9 | 28 | 1.2 |
| Sexual abuse | 64 | 2.4 | 62 | 2.6 |
| Physical neglect | 541 | 19.9 | 421 | 17.5 |
| Emotional neglect | 67 | 2.5 | 51 | 2.1 |
| Bullying | 839 | 30.8 | 647 | 27.0 |
| Winter birth | 872 | 32.1 | 699 | 29.1 |
| Hearing impairment | 223 | 8.2 | 142 | 5.9 |
| Cannabis use | 5 | 0.2 | 3 | 0.1 |

| At outcome assessment (3-year follow-up) | Mean | SD | Mean | SD |
| --- | --- | --- | --- | --- |
| Age (years) | 12.9 | 0.7 | 13.0 | 0.7 |
| Distressing PEs | n | % | n | % |
| Past-month | 209 | 7.7 | 352 | 14.7 |
| Lifetime ($\geq 1$ wave) | 850 | 31.2 | 901 | 37.5 |
| Repeating $\geq 2$ waves | 333 | 12.2 | 408 | 17.0 |
| Repeating $\geq 3$ waves | 130 | 4.8 | 159 | 6.6 |
| Persisting (all 4 waves) | 34 | 1.3 | 64 | 2.7 |

### Association of PRS-SCZ on PEs

For females, PRS-SCZ_75_ was not significantly associated with past-month distressing PEs but with lifetime (OR 1.47 [95%CI 1.12, 1.92]) and repeating (≥2 waves) distressing PEs (OR 1.45 [95%CI 1.05, 2.00]). However, no significant associations were found for repeating (≥ 3 waves) and persisting distressing PEs (Table 2, Model 1, Fig. 1a). Findings from Model 2, additionally adjusted for family income and parental education, revealed similar results (Table 2).

**Fig. 1.**
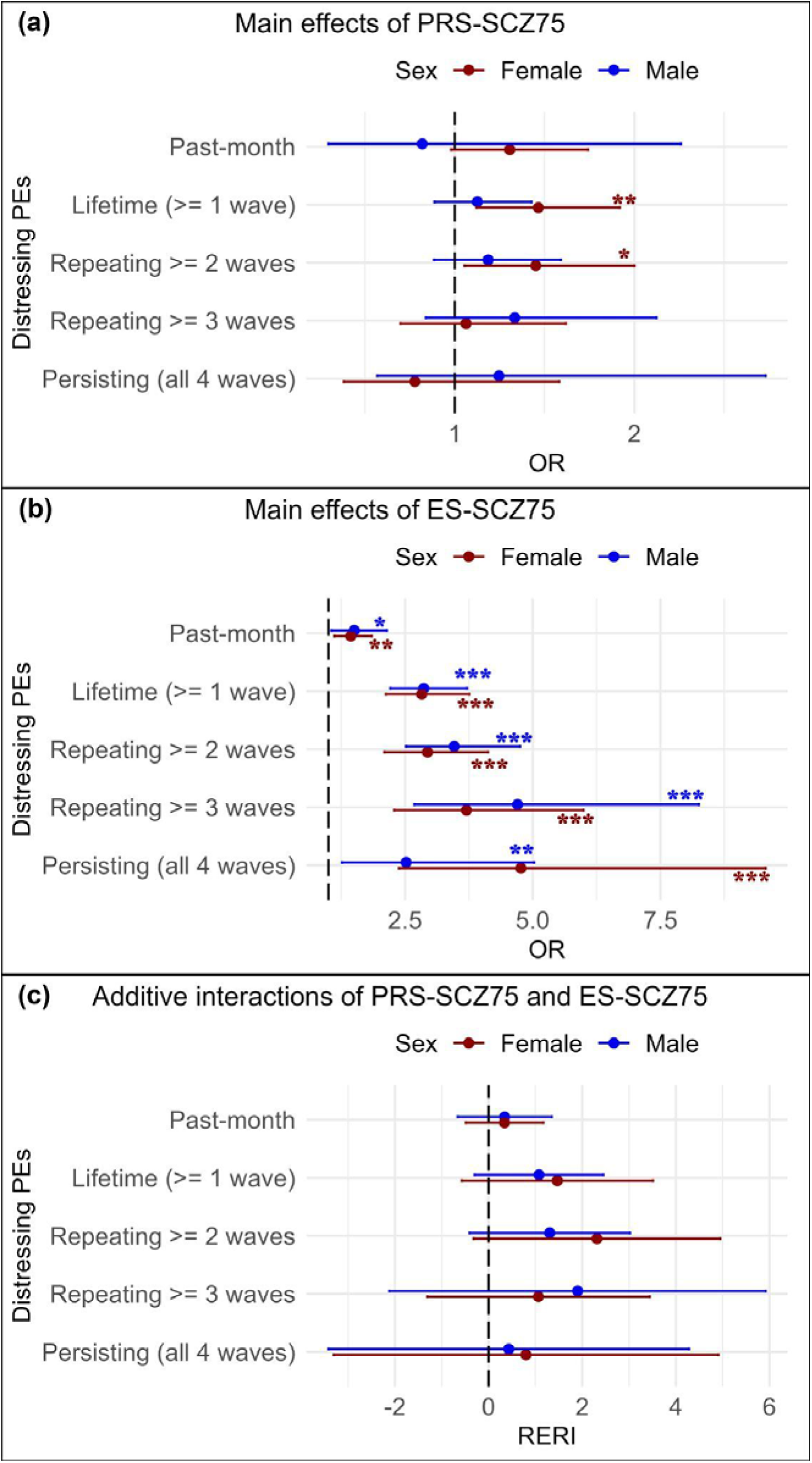
Visualization of age-adjusted main effects of PRS-SCZ_75_ (a) and ES-SCZ_75_ (b) and their joint effects (c) on distressing PEs in male and female adolescents (* p < .05, ** p < .01, *** p < .001)

**Table 2.** Main associations of PRS-SCZ_75_ with distressing PEs at 3-year follow-up in male and female adolescents.

| Distressing<br>PEs | Males |  |  |  |  |  | Females |  |  |  |  |  |
| --- | --- | --- | --- | --- | --- | --- | --- | --- | --- | --- | --- | --- |
|  | Model 1<sup>a</sup> (N = 2,721) |  |  | Model 2<sup>b</sup> (N = 2,602) |  |  | Model 1<sup>a</sup> (N = 2,401) |  |  | Model 2<sup>b</sup> (N = 2,310) |  |  |
|  | OR | 95% CI | *p*-value | OR | 95% CI | *p*-value | OR | 95% CI | *p*-value | OR | 95% CI | *p*-value |
| Past-month | 0.82 | 0.30 to 2.26 | .700 | 0.67 | 0.23 to 2.02 | .480 | 1.31 | 0.98 to 1.74 | .068 | 1.28 | 0.95 to 1.73 | .098 |
| Lifetime<br>(≥ 1 wave) | 1.13 | 0.89 to 1.43 | .329 | 1.08 | 0.85 to 1.37 | .530 | 1.47 | 1.12 to 1.92 | .005* | 1.39 | 1.07 to 1.81 | .015* |
| Repeating<br>≥ 2 waves | 1.19 | 0.88 to 1.59 | .254 | 1.12 | 0.84 to 1.51 | .445 | 1.45 | 1.05 to 2.00 | .023* | 1.42 | 1.02 to 1.97 | .038* |
| Repeating<br>≥ 3 waves | 1.33 | 0.84 to 2.12 | .223 | 1.31 | 0.80 to 2.16 | .282 | 1.06 | 0.70 to 1.62 | .776 | 0.99 | 0.64 to 1.53 | .968 |
| Persisting<br>(all 4 waves) | 1.25 | 0.57 to 2.73 | .582 | 1.19 | 0.54 to 2.64 | .665 | 0.78 | 0.38 to 1.58 | .488 | 0.62 | 0.27 to 1.40 | .252 |

Note: PEs, psychotic experiences; OR, odds ratio; CI, confidence interval
<sup>a</sup> Model 1: Adjusted for age; <sup>b</sup> Model 2: Adjusted for age, family income, and parental education
\* *p*-values <.05

For males, PRS-SCZ_75_ was not significantly associated with past-month distressing PEs or any secondary outcomes (Table 2, Model 1, Fig. 1a). The results from Model 2 further supported these findings (Table 2).

Sensitivity analyses of an unrelated sample confirmed a differential association pattern between PRS-SCZ_75_ and lifetime and repeating (≥2 waves) distressing PEs in females but not males (Online Resource 2). Follow-up analyses indicated a statistically significant sex difference for lifetime distressing PEs (≥1 wave) across both adjusted models (p<.001). No other significant sex differences were observed in the association between PRS-SCZ_75_ and PEs.

### Association of ES-SCZ on PEs

For females, ES-SCZ_75_ was significantly associated with past-month distressing PEs (OR 1.44 [95%CI 1.12, 1.85]) and all secondary outcomes (lifetime: OR 2.83 [95%CI 2.13, 3.76]; repeating ≥2 waves: OR 2.94 [95%CI 2.10, 4.12]; repeating ≥3 waves: OR 3.70 [95%CI 2.29, 5.99]; and persisting distressing PEs: OR 4.77 [95%CI 2.38, 9.56]). As shown in Fig. 1b, the ORs of ES-SCZ_75_ generally increase for more PE persistence. The results from Model 2 further supported these findings (Table 3).

**Table 3.** Main associations of ES-SCZ_75_ at 2-year follow-up with distressing PEs at 3-year follow-up in male and female adolescents.

| Distressing<br>PEs | Males |  |  |  |  |  | Females |  |  |  |  |  |
| --- | --- | --- | --- | --- | --- | --- | --- | --- | --- | --- | --- | --- |
|  | Model 1<sup>a</sup> (N = 2,721) |  |  | Model 2<sup>b</sup> (N = 2,602) |  |  | Model 1<sup>a</sup> (N = 2,401) |  |  | Model 2<sup>b</sup> (N = 2,310) |  |  |
|  | OR | 95% CI | *p*-value | OR | 95% CI | *p*-value | OR | 95% CI | *p*-value | OR | 95% CI | *p*-value |
| Past-month<sup>c</sup> | 1.50 | 1.06 to 2.14 | .023<sup>*</sup> | 1.47 | 1.00 to 2.15 | .050 | 1.44 | 1.12 to 1.85 | .005<sup>*</sup> | 1.35 | 1.03 to 1.76 | .031<sup>*</sup> |
| Lifetime<br>(≥ 1 wave) | 2.87 | 2.21 to 3.71 | <.001<sup>*</sup> | 2.70 | 2.09 to 3.49 | <.001<sup>*</sup> | 2.83 | 2.13 to 3.76 | <.001<sup>*</sup> | 2.48 | 1.87 to 3.28 | <.001<sup>*</sup> |
| Repeating<br>≥ 2 waves | 3.46 | 2.52 to 4.75 | <.001<sup>*</sup> | 3.23 | 2.38 to 4.39 | <.001<sup>*</sup> | 2.94 | 2.10 to 4.12 | <.001<sup>*</sup> | 2.62 | 1.87 to 3.66 | <.001<sup>*</sup> |
| Repeating<br>≥ 3 waves | 4.70 | 2.68 to 8.25 | <.001<sup>*</sup> | 4.35 | 2.39 to 7.92 | <.001<sup>*</sup> | 3.70 | 2.29 to 5.99 | <.001<sup>*</sup> | 3.15 | 1.97 to 5.04 | <.001<sup>*</sup> |
| Persisting<br>(all 4 waves) | 2.52 | 1.27 to 5.03 | .009<sup>*</sup> | 2.41 | 1.16 to 5.03 | .019<sup>*</sup> | 4.77 | 2.38 to 9.56 | <.001<sup>*</sup> | 4.08 | 2.01 to 8.29 | <.001<sup>*</sup> |

Note: PEs, psychotic experiences; OR, odds ratio; CI, confidence interval
<sup>a</sup> Model 1: Adjusted for age; <sup>b</sup> Model 2: Adjusted for age, family income, and parental education
<sup>c</sup> Additionally adjusted for distressing PEs up to 2-year-follow-up
<sup>\*</sup> *p*-values <.05

For males, ES-SCZ_75_ was significantly associated with past-month distressing PEs (OR 1.50 [95%CI 1.06, 2.14]) and all secondary outcomes (lifetime: OR 2.87 [95%CI 2.21, 3.71]; repeating ≥2 waves: OR 3.46 [95%CI 2.52, 4.75]; repeating ≥3 waves: OR 4.70 [95%CI 2.68, 8.25]; and persisting distressing PEs: OR 2.52 [95%CI 1.27, 5.03]). As shown in Fig. 1b, the ORs of ES-SCZ_75_ generally increase for more PE persistence, except for persisting (4 waves) distressing PEs. The results from Model 2 further supported these findings (Table 3).

Sensitivity analyses of an unrelated sample revealed a similar pattern of increasing ORs for ES-SCZ_75_ for more PE persistence in females and males. No significant sex differences in the ORs of ES-SCZ_75_ were observed for distressing PEs of any definition (Online Resource 2).

### Joint interaction of PRS-SCZ and ES-SCZ on PEs

When considered jointly, the isolated genetic risk state (PRS-SCZ_75_=1 & ES-SCZ_75_=0) was not associated with past-month distressing PEs in either females (Table 4) or males (Table 5). However, the isolated exposomic risk state (PRS-SCZ_75_=0 & ES-SCZ_75_=1) increased the odds of having past-month distressing PEs at a statistically significant level in females (OR 1.37 [95% CI 1.02, 1.85]), but only at a trend level in males (OR 1.45 [95% CI 0.98, 2.16]). Similarly, the combined risk state (PRS-SCZ_75_=1 & ES-SCZ_75_=1) was associated with past-month distressing PEs significantly in females (OR 1.82 [95% CI 1.19, 2.77]) and showed a trend-level association in males (OR 1.68 [95% CI 0.97, 2.90]). However, the combined effect did not significantly differ from the sum of the ORs of having either risk state alone, indicating a null additive interaction between PRS-SCZ_75_ and ES-SCZ_75_ in either sex (Table 4-5, Fig.1c).

**Table 4.**
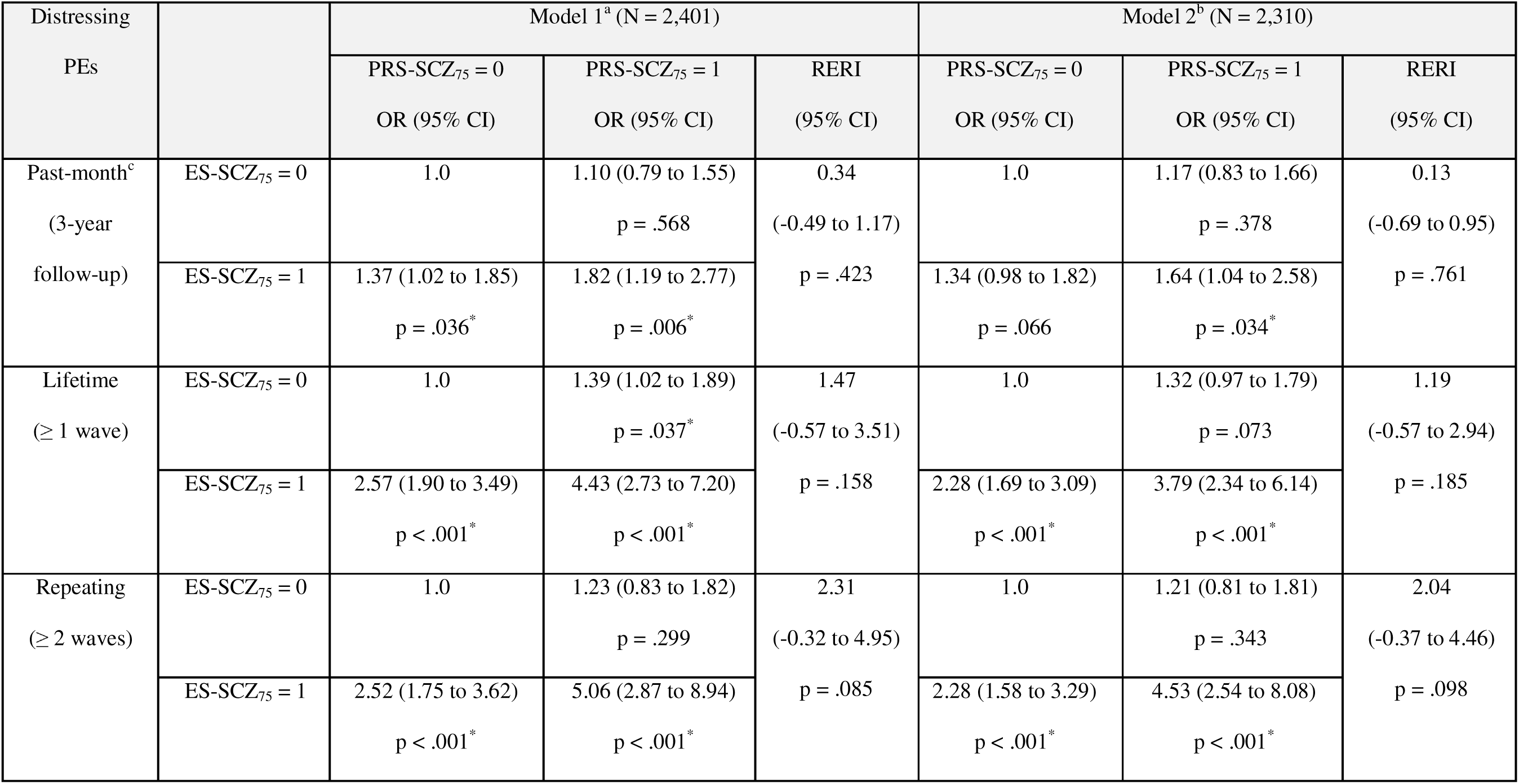
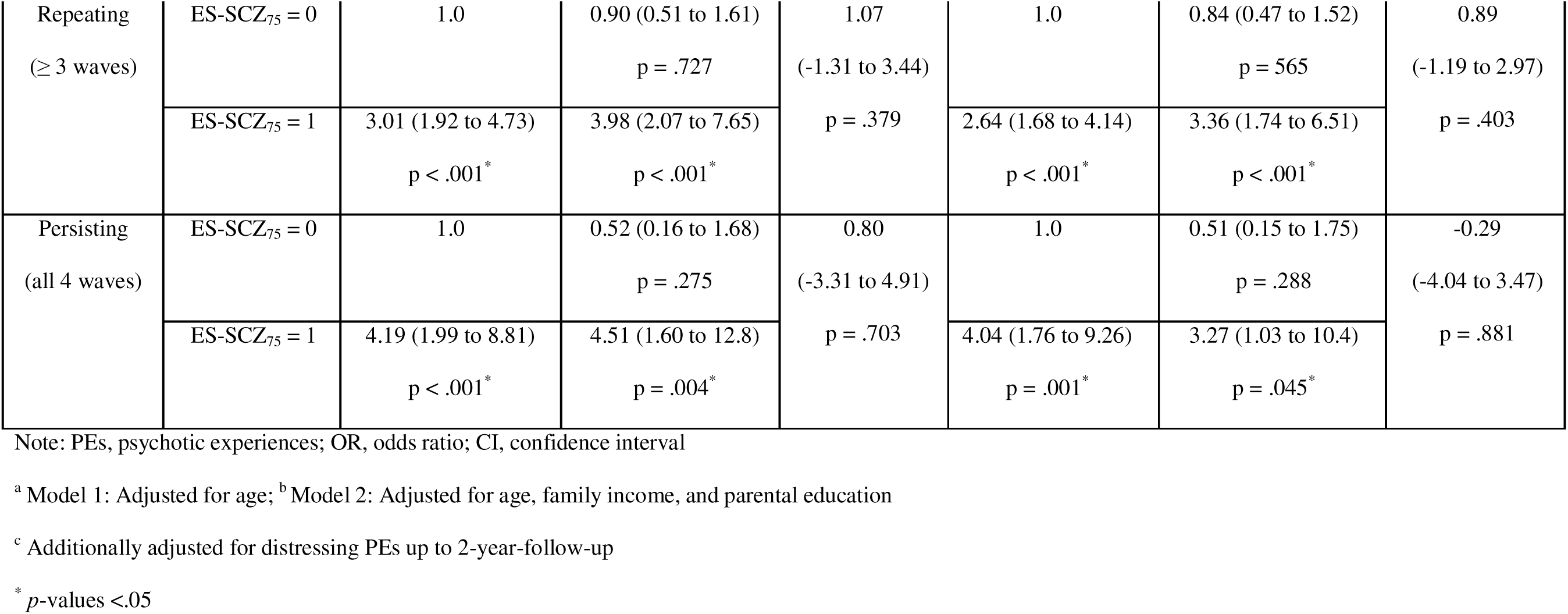
Joint associations of PRS-SCZ_75_ and ES-SCZ_75_ at 2-year follow-up with distressing PEs at 3-year follow-up in female adolescents.

**Table 5.** Joint associations of PRS-SCZ_75_ and ES-SCZ_75_ at 2-year follow-up with distressing PEs at 3-year follow-up in male adolescents.

| Distressing PEs |  | Model 1<sup>a</sup> (N = 2,721) |  |  | Model 2<sup>b</sup> (N = 2,602) |  |  |
| --- | --- | --- | --- | --- | --- | --- | --- |
|  |  | PRS-SCZ<sub>75</sub> = 0<br>OR (95% CI) | PRS-SCZ<sub>75</sub> = 1<br>OR (95% CI) | RERI<br>(95% CI) | PRS-SCZ<sub>75</sub> = 0<br>OR (95% CI) | PRS-SCZ<sub>75</sub> = 1<br>OR (95% CI) | RERI<br>(95% CI) |
| Past-month<sup>c</sup><br>(3-year follow-up) | ES-SCZ<sub>75</sub> = 0 | 1.0 | 0.88 (0.52 to 1.49)<br>p = .638 | 0.34<br>(-0.66 to 1.35)<br>p = .503 | 1.0 | 0.82 (0.46 to 1.44)<br>p = .485 | 0.37<br>(-0.66 to 1.40)<br>p = .482 |
|  | ES-SCZ<sub>75</sub> = 1 | 1.45 (0.98 to 2.16)<br>p = .065 | 1.68 (0.97 to 2.90)<br>p = .063 |  | 1.39 (0.91 to 2.14)<br>p = .129 | 1.58 (0.87 to 2.87)<br>p = .133 |  |
| Lifetime<br>(≥ 1 wave) | ES-SCZ<sub>75</sub> = 0 | 1.0 | 0.98 (0.72 to 1.32)<br>p = .880 | 1.08<br>(-0.30 to 2.45)<br>p = .125 | 1.0 | 0.94 (0.69 to 1.26)<br>p = .667 | 0.91<br>(-0.34 to 2.16)<br>p = .153 |
|  | ES-SCZ<sub>75</sub> = 1 | 2.55 (1.93 to 3.37)<br>p < .001* | 3.61 (2.41 to 5.41)<br>p < .001* |  | 2.42 (1.83 to 3.19)<br>p < .001* | 3.27 (2.18 to 4.89)<br>p < .001* |  |
| Repeating<br>(≥ 2 waves) | ES-SCZ<sub>75</sub> = 0 | 1.0 | 0.97 (0.64 to 1.47)<br>p = .882 | 1.31<br>(-0.41 to 3.02)<br>p = .136 | 1.0 | 0.91 (0.60 to 1.40)<br>p = .675 | 1.11<br>(-0.43 to 2.66)<br>p = .157 |
|  | ES-SCZ<sub>75</sub> = 1 | 3.08 (2.18 to 4.35)<br>p < .001* | 4.35 (2.79 to 6.78)<br>p < .001* |  | 2.90 (2.07 to 4.07)<br>p < .001* | 3.93 (2.55 to 6.05)<br>p < .001* |  |
| Repeating<br>(≥ 3 waves) | ES-SCZ<sub>75</sub> = 0 | 1.0 | 1.16 (0.57 to 2.36)<br>p = .673 | 1.90<br>(-2.12 to 5.92)<br>p = .354 | 1.0 | 1.10 (0.52 to 2.32)<br>p = .803 | 1.91<br>(-2.05 to 5.86)<br>p = .345 |
|  | ES-SCZ<sub>75</sub> = 1 | 4.39 (2.38 to 8.08)<br>p < .001<sup>*</sup> | 6.45 (2.95 to 14.1)<br>p < .001<sup>*</sup> |  | 3.94 (2.08 to 7.49)<br>p < .001<sup>*</sup> | 5.95 (2.61 to 13.6)<br>p < .001<sup>*</sup> |  |
| Persisting<br>(all 4 waves) | ES-SCZ<sub>75</sub> = 0 | 1.0 | 1.32 (0.44 to 3.93)<br>p = .615 | 0.43<br>(-3.42 to 4.28)<br>p = .826 | 1.0 | 1.28 (0.43 to 3.83)<br>p = .658 | 0.27<br>(-3.31 to 3.86)<br>p = .881 |
|  | ES-SCZ<sub>75</sub> = 1 | 2.74 (1.13 to 6.66)<br>p = .026<sup>*</sup> | 3.49 (1.14 to 10.7)<br>p = .028<sup>*</sup> |  | 2.60 (1.05 to 6.44)<br>p = .038<sup>*</sup> | 3.16 (1.01 to 9.91)<br>p = .049<sup>*</sup> |  |

Note: PEs, psychotic experiences; OR, odds ratio; CI, confidence interval
<sup>a</sup> Model 1: Adjusted for age; <sup>b</sup> Model 2: Adjusted for age, family income, and parental education
<sup>c</sup> Additionally adjusted for distressing PEs up to 2-year-follow-up
<sup>\*</sup> *p*-values <.05

For secondary outcomes, the isolated genetic risk state was significantly associated with lifetime distressing PEs (OR 1.39 [95% CI 1.02, 1.89]) only in females. No significant associations were observed for other secondary outcomes in either sex. The isolated exposomic risk state and the combined risk state were associated with all secondary outcomes in both sexes (Table 4-5). However, no significant additive interactions were detected (Table 4-5, Fig. 1c). These findings were consistent in both adjusted models (Table 4-5) and were further confirmed by the sensitivity analyses of an unrelated sample (Online Resource 2).

## Discussion

This study investigated the joint and independent associations of polygenic and exposomic liabilities for schizophrenia with distressing PEs and their persistence in male and female adolescents. PRS-SCZ_75_ was significantly associated with lifetime (≥1 wave) and repeating (≥2 wave) distressing PEs in females but not with any PE definition in males. ES-SCZ_75_ was associated with all PE definitions with a trend of higher ORs for more PE persistence in both sexes. No significant additive interactions between PRS-SCZ_75_ and ES-SCZ_75_ were observed for any PE definition.

Our finding that PRS-SCZ was significantly associated with distressing PEs only in females suggests a sex-dependent influence of genetic risks on subclinical psychosis in early adolescence. This contradicts evidence from adults showing more dominant PRS-SCZ influences on psychosis-related phenotypes, including poor cognitive performance and corresponding brain function, in men(Koch et al. 2022; Koch et al. 2021; Mitjans et al. 2024). Combined with poorer cognitive function and more pronounced negative symptoms reported in men with schizophrenia(Giordano et al. 2021), such evidence implies that males are more susceptible to neurocognitive impairments, particularly under genetic risk for schizophrenia. Familial aggregation and CNV studies, however, indicate that women with schizophrenia carry a higher genetic risk burden than men(Goldstein et al. 1990; Han et al. 2016). This pattern aligns with the ‘female protective effect’ hypothesis, which posits that females are inherently more resilient to certain neurodevelopmental and psychiatric conditions, like schizophrenia, disproportionately affecting men. Therefore, females may require a greater genetic load to surpass the threshold for developing psychosis-related phenotypes(Jacquemont et al. 2014; Robinson et al. 2013). Under this framework, our finding on female-specific association between PRS-SCZ and distressing PEs might be an indirect signal of this protective effect rather than evidence of a stronger genetic influence in females. Specifically, if males are more vulnerable to schizophrenia-related traits even at lower genetic risk levels, their psychosis-related phenotypes may emerge through other phenotypes (cognitive deficits or negative symptoms) rather than distressing PEs. In contrast, the emergence of distressing PEs in females might indicate that only those with a higher genetic load for schizophrenia surpass the threshold for experiencing such symptoms. Interestingly, some studies reported a male-specific association of PRS-SCZ with trait-like psychosis-related phenotypes, such as schizotypy and negative symptoms, but not with state-like positive symptoms such as PEs(Docherty et al. 2020; Mas-Bermejo et al. 2024). Combined with our findings, this suggests that genetic risks for schizophrenia may have sex-dependent effects, with men more likely to manifest stable, trait-like phenotypes (e.g., schizotypy, negative symptoms, and cognitive deficits), while women may be more prone to developing transient, state-like symptoms such as distressing PEs. However, due to methodological differences among studies (e.g., the age of the studied population, genetic risk markers, and study design), future research is still needed to validate our proposed hypothesis.

From a transdiagnostic perspective, it is important to note that distressing PEs are not specific to psychotic disorders. A meta-analysis has shown that the pooled OR of PEs for affective disorders (3.83) in childhood and adolescence is comparable to that for psychotic disorders (3.96)(Healy et al. 2019). Given that female preponderance in depression emerges around age 12(Salk et al. 2017), the female-specific PRS-SCZ association at age 13 in our study may reflect a general increase in mental ill-health rather than an association specific to PEs. Moreover, prior studies(Stainton et al. 2021; Zammit et al. 2013), like ours, found that female adolescents report higher levels of distress and recurring PEs than males, rendering the analytical power for females stronger than males. Interestingly, evidence from a general population sample reported a higher rate of positive PEs that was confounded by depressive symptoms in females(Maric et al. 2003). Therefore, large prospective studies assessing PEs and associated psychopathology across the development are needed to clarify these findings.

Relative to PRS-SCZ, sex-stratified analyses of the influences of ES-SCZ are even more scarce. In this study, no statistically significant sex differences were detected for any outcomes. However, the ORs for persisting distressing PEs (4 waves) appeared almost double in females. Indeed, a stronger influence of ES-SCZ in women was reported for physical health, suggesting a greater sensitivity to the environmental insults composing ES-SCZ among females(Paquin et al. 2023). Interestingly, we consistently observed significant associations of ES-SCZ, but not PRS-SCZ, with all outcomes and a dose-response relationship with PE persistence in both sexes, highlighting a universally more dominant impact of the environmental risk than the genetic risk for schizophrenia on distressing PEs. Concordantly, evidence from a large male schizophrenia cohort showed a substantial impact of cumulative environmental risk, but not schizophrenia polygenic risk, on the age at schizophrenia onset(Stepniak et al. 2014). However, the limited research on sex differences calls for more comparative analysis of genetic and environmental contributions to confirm our findings.

Apart from the independent associations, prior evidence has shown that the influences of genomic and exposomic risk for schizophrenia are synergistic(Pries et al. 2020). Our prior unstratified analyses of the ABCD dataset also found significant additive interactions between ES-SCZ_75_ and PRS-SCZ_75_ for distressing PEs recurring in ≥1-2 waves(Di Vincenzo et al. 2025). However, the current sex-stratified analyses, halving the sample size, could not detect significant additive interactions, although the ORs of the combined risk state tended to be greater than either risk alone for most outcomes (Table 4-5). Considering that the CIs of the RERIs are wide and the observed RERIs are above zero in both sexes (see Fig. 1c), the non-significant additive interaction could reflect a type II error of our sex-stratified analysis.

### Strengths and limitations

To our knowledge, this is the first study to examine sex differences in how an established aggregated environmental risk score for schizophrenia, independently and with polygenic risk, influences PEs in early adolescents. Focusing on distressing and persistent PEs enhances the clinical relevance of our findings, as both are key predictors of poor outcomes(Karcher et al. 2020; Staines et al. 2023). Nevertheless, the findings need to be considered in light of limitations.

First, we restricted the analysis to the European subsample to ensure good performance of PRS-SCZ based on recent GWAS, limiting generalizability to other populations. Second, PEs in the ABCD dataset were self-reported, which may lead to false positives compared to interview-based assessments(Staines et al. 2023). However, prior evidence has shown that, although to a lesser extent, non-validated self-reported PEs are significantly associated with psychopathology similar to clinically validated PEs and attenuated psychosis syndrome(Moriyama et al. 2019). Furthermore, even false-positive self-reported PEs confirmed by a clinical interview appear to be associated with later mental health problems(van der Steen et al. 2019), supporting the clinical significance of our findings. Third, while genomic risks always precede outcomes, exposomic risks may have a bidirectional relationship with distressing PEs, potentially inflating associations. To mitigate this, we adjusted for prior-wave distressing PEs in past-month analyses, reducing the likelihood of overestimating ES-SCZ_75_ effects. However, this approach could not be applied to secondary outcomes. Lastly, our sex-stratified analyses have limited the sample, increasing the chances of type II errors for low prevalent outcomes (e.g., persisting distressing PEs) or potentially weak associations (e.g., interaction effects). This might be particularly relevant for males, given their lower prevalence of distressing PEs.

## Conclusion

Our findings on the association of PRS-SCZ and distressing PEs only in females underscore the importance of a sex-sensitive approach in studying genetic contribution to subclinical psychosis. Dose-response effects of ES-SCZ on PE persistence in both males and females highlight environmental risks as universal targets to prevent PEs and associated psychopathology in early adolescence.

## Supporting information

Online Resource 1

Online Resource 2

## Ethical Approval

The ABCD study was approved by the Centralized Institutional Review Board (IRB) of the University of California-San Diego and local research site IRBs in accordance with their IRB-approved protocols, state regulations, and local resources.

## Consent to Participate

As a part of the ABCD study, written informed consent and assent were derived from participating parents/caregivers and adolescents, respectively.

## Consent to Publish

All authors transfer, assign or otherwise convey all copyright ownership to Archives of Women’s Mental Health in the event the manuscript is published.

## Author Contributions

S.G. and L.K.P. contributed equally as co-last authors. They, along with T.P., designed the study. B.D.L. and A.A.M. prepared the genetic data, while T.P., under S.G. and L.K.P.’s supervision, processed and analyzed it. T.P. and L.K.P. drafted the first version of the manuscript. S.G. provided critical revision of the manuscript for important intellectual content. J.V.O., B.P.F.R., and S.G. secured funding. All authors reviewed, edited and approved the final manuscript.

## Statements and Declarations

J. van Os and S. Guloksuz are supported by the Ophelia research project, ZonMw grant 636340001. B. Rutten was funded by a Vidi award (91718336) from the Netherlands Scientific Organisation. J. van Os, S. Guloksuz, B. Rutten, L. K. Pries, B. D. Lin, and A. G. Arias Magnasco are supported by the YOUTH-GEMs project, funded by the European Union’s Horizon Europe program under the grant agreement number: 101057182. T. Prachason is supported by the Faculty of Medicine Ramathibodi Hospital’s Research Talent Grant.

## Data availability

Data used in the preparation of this article were obtained from publicly available data from the Adolescent Brain Cognitive Development Study (https://abcdstudy.org), held in the National Institute of Mental Health Data Archive.

