## Supplementary material for "Sex Differences in the Independent and Combined Effects of Genomic and Exposomic Risks for Schizophrenia on Distressing Psychotic Experiences: Insights from the ABCD Study": Online Resource 1

**Journal:** Archives of Women’s Mental Health

Thanavadee Prachason^1^, Angelo Arias-Magnasco^2^, Bochao Danae Lin^2^, Jim van Os^2-4^, Bart P.F. Rutten^2^, Lotta-Katrin Pries^2^* and Sinan Guloksuz^2,5^*^#^

*^1^Department of Psychiatry, Faculty of Medicine Ramathibodi Hospital, Mahidol University, Bangkok, Thailand.*

*^2^Department of Psychiatry and Neuropsychology, School for Mental Health and Neuroscience, Maastricht University Medical Centre, Maastricht, The Netherlands.*

*^3^Department of Psychiatry, UMC Utrecht Brain Centre, University Medical Centre Utrecht, Utrecht University, Utrecht, The Netherlands.*

*^4^Department of Psychosis Studies, Institute of Psychiatry, Psychology & Neuroscience, King's College London, London, UK.*

*^5^Department of Psychiatry, Yale University School of Medicine, New Haven, Connecticut.*

* Shared last author

**^#^ Corresponding author:** Sinan Guloksuz, Department of Psychiatry and Neuropsychology, School for Mental Health and Neuroscience, Maastricht University Medical Center, P.O. Box 616 6200 MD Maastricht, The Netherlands; tel: 31-433-88-4071, fax: 31433-88-4122,

### **Supplementary measurements**

#### *Exposome score for schizophrenia (ES-SCZ)*

Lifetime environmental exposures obtained from ABCD Study dataset were recoded as binary variables (0 = “absent”; 1 = “present”) in order to generate ES-SCZ^1^. The following information describes the procedure of dichotomization in detail.

*Emotional neglect*. Emotional neglect was derived from five items of the *Child Report of Parent Behavior Inventory* (CRPBI)^2^. Children reported perceived levels of parents/primary caregivers’ warmth and closeness across a 3-point Likert scale (1 = “Not like him/her”; 2 = “Somewhat like him/her”; 3 = “A lot like him/her”). Similar to Stinson et al.^3^, emotional neglect was regarded as present if children answered “Not like him/her” in at least two out of five items of the CRPBI.

*Physical neglect.* Consistent with previous research^3,4^, physical neglect was considered present if any of the conditions below had been experienced by youth. Food shortage was reported in the *Parent Demographics Survey* and in the *Longitudinal Parent Demographics Survey* (0 = “No”; 1 = “Yes”). History of maternal or paternal alcohol/substance use that had caused work, legal, or social problems, or required a treatment program was reported by parents in two items of the *Family History Assessment Part 1* (0 = “No”; 1 = “Yes”). Poor parental supervision was considered positive if “Never” or “Almost never” were answered by youth in any of the three items on the *Parental Monitoring Survey*.

*Emotional abuse.* Following Stinson et al.^3^, emotional abuse was considered present if parents answered “Yes” to the item “A family member threatened to kill your child” on the PTSD module of the *Kiddie Schedule for Affective Disorders and Schizophrenia for DSM-5 ^5,6^*.

*Physical abuse*. Consistent with previous research^3^, physical abuse was considered endorsed when parents answered “Yes” to any of the following items on the *Kiddie Schedule for Affective Disorders and Schizophrenia for DSM-5*: “Shot, stabbed, or beaten brutally by a grown up in the home” or “Beaten to the point of having bruises by a grown up in the home”.

*Sexual abuse.* When parents endorsed any of the following items on the *Kiddie Schedule for Affective Disorders and Schizophrenia for DSM-5*: “A grown up in the home touched your child in their privates, had your child touch their privates, or did other sexual things to your child”, “An adult outside your family touched your child in their privates, had your child touch their privates or did other sexual things to your child”, or “A peer forced your child to do something sexually”, sexual abuse was considered present^3^.

*Cannabis use.* Lifetime cannabis use was regarded positive (coded as “1”) if any of the following condition were met. Having ever used cannabis at least 10 times was reported by youth on the *ABCD Youth Substance Use Interview* at baseline. Use of cannabis at least 10 times during the previous year was reported by youth on the *ABCD Youth Substance Use Introduction and Patterns* at the yearly follow-up, and on *the ABCD Timeline Follow-back Survey Calendar Scores (TLFB)* across the third and fourth waves of follow-up. Use of cannabis 5 times or more during the previous 6 months was reported on the *ABCD Youth Marijuana Illicit Drug Measures* at the mid-year follow-up. Having ever experienced craving, withdrawal symptoms, or tolerance was reported on the *ABCD Parent Diagnostic Interview for DSM-5 (KSADS) Drug Use Disorder Individual Questions*.

*Winter birth.* Winter birth was derived by leveraging the precise date of the interview at baseline and the child’s age in months, both available in the *Demographics Survey*. The variable was coded as positive (“1”) if birthday fell in December, January, February, or March.

*Hearing impairment*. This variable was considered endorsed when parents answered “Yes” at least once to the questions: “Has she/he ever been to a doctor for any of these things… Hearing Problem” on the *ABCD Parent Medical History Questionnaire* at baseline, and: “Since we last saw you, has she/he been to a doctor for any of these things? Hearing Problem” on the *ABCD Longitudinal Parent Medical History Questionnaire* at follow-up.

*Bullying.* Bullying was considered endorsed when parents answered “Yes” to the question: “Does your child have any problems with bullying at school or in your neighborhood?” on the *ABCD Parent Diagnostic Interview for DSM-5 Background Items Full* at baseline, and/or to the question: “Since we last saw you, did your child have any problems with bullying at school or in your neighbourhood?” on the *ABCD Longitudinal Parent Diagnostic Interview for DSM-5 Background Items Full (KSAD)* at follow-up.

Similar to the previous literature^1^, ES-SCZ was calculated by summing the nine exposures multiplied by their log odds (i.e. weighted risks) for schizophrenia and added by 2 for ease of interpretation as in the following formula: ES-SCZ=(cannabis use*1.31) + (winter birth*0.03) + (hearing impairment*1.18) + (emotional abuse*0.78) + (physical abuse*-0.39) + (sexual abuse*0.86) + (emotional neglect*0.44) + (physical neglect*0.25) + (bullying*1.35) + 2

*Covariates*

Baseline family income with following levels: 1 = Less than $5,000; 2 = $5,000 through $11,999; 3 = $12,000 through $15,999; 4 = $16,000 through $24,999; 5 = $25,000 through $34,999; 6 = $35,000 through $49,999; 7 = $50,000 through $74,999; 8 = $75,000 through $99,999; 9 = $100,000 through $199,999; 10 = $200,000 and greater

Highest parental education with following levels: 0 = Never attended/Kindergarten only; 1 = 1st grade; 2 = 2nd grade; 3 = 3rd grade; 4 = 4th grade; 5 = 5th grade; 6 = 6th grade; 7 = 7th grade; 8 = 8th grade; 9 = 9th grade; 10 = 10th grade; 11 = 11th grade; 12 = 12th grade; 13 = High school graduate; 14 = GED or equivalent; 15 = Some college; 16 = Associate degree: Occupational; 17 = Associate degree: Academic Program; 18 = Bachelor's degree (ex. BA); 19 = Master's degree (ex. MA); 20 = Professional School degree (ex. MD); 21 = Doctoral degree (ex. PhD)

#### **Table S1** The number of missing data in European samples

| Variables | Number of missing data | |
| --- | --- | --- |
|  | Males (n=2,984) | Females (n=2,665) |
| Primary outcome (at 3-year follow-up) |  |  |
| Psychotic experiences | 260 | 262 |
| Lifetime exposure (up to 2-year follow-up) |  |  |
| Physical abuse | 1 | 5 |
| Emotional abuse | 1 | 5 |
| Sexual abuse | 1 | 5 |
| Physical neglect | 0 | 0 |
| Emotional neglect | 2 | 1 |
| Bullying | 0 | 0 |
| Winter birth | 1 | 0 |
| Hearing impairment | 0 | 1 |
| Cannabis use | 1 | 0 |
| Covariates |  |  |
| Age (at 3-year follow-up) | 258 | 256 |
| Family income | 0 | 0 |
| Parental education | 0 | 0 |

#### *Polygenic risk score for schizophrenia (PRS-SCZ)*

Quality Control, Imputation and Polygenic Risk Score calculations - ABCD Genotype Data Release 4.0

The ABCD sample consisted in 11,101 individuals from which genomic DNA was extracted from buccal mucosa using Chemagic STAR DNA Saliva 4k Kit (Hamilton Robotics, Reno, NV, United States). Rutger University Cell and DNA Repository (RUCDR) performed a genome-wide genotyping using the Affymetrix NIDA SmokeScreen Array (Affymetrix, Santa Clara, CA, USA), resulting in the genotyping of 733,329 Single-Nucleotide Polymorphisms (SNPs). The genotyped Quality Controlled (QCed) data was in binary PLINK format and contained 516,598 genetic variants referenced in positive strand and aligned to GRCh37 (hg19). Further details can be found in the NDA 4.0 Release Notes – Genetics (<https://nda.nih.gov/study.html?id=1299>).

**1. Pre-imputation Quality Control**

Pre-imputation Quality Control (QC) was performed using PLINK 1.9^9^. The initial phase involved genotypic QC, which proceeded as follows: i. RUCDR performed DNA quality controls based on calling signals and variant call rates. ii. ABCD DAIRC performed the subsequent study-based QC process, following the recommendation of Ricopili pipeline (<https://nda.nih.gov/study.html?id=1299>).

Following the genotype QC, a strict SNP QC was performed. This process was divided into two phases: SNP QC and sample QC. The SNP QC consisted of the following steps: (1) removing SNPs with a Minor Allele Frequency (MAF) < 10% (2) and a Hardy-Weinberg Equilibrium p-value < 1e-5, (3) excluding SNPs located in long Linkage Disequilibrium (LD) regions, (4) removing non-autosomal SNPs, (5) pruning SNPs on maximum LD r^2^ = 0.5. The Sample QC consisted of (6) removing bad QC samples, (7) removing samples which were mismatched between genetic sex and reported sex, (8) removing samples with problematic genotyping batch, and (9) heterozygosity outliers (+/- 2 Standard Deviation (SD) samples. The detail of QC steps is shown in Table S3.

#### **Table S2** Pre-imputation QC steps.

| Step | SNP start | SNP end | Subjects start | Subjects end |
| --- | --- | --- | --- | --- |
| Strict SNP QC for removal of bad samples – remove SNPs with MAF < 10% and HWE 1e-5 | 516,598 | 230,733 | 11,101 | 11,101 |
| Strict SNP QC – Remove SNPs in long LD regions | 230,733 | 223,421 | 11,101 | 11,101 |
| Strict SNP QC – Remove insertions and deletions | 223,421 | 222,422 | 11,101 | 11,101 |
| Strict SNP QC – Only autosomes (Chr 1 – 22) | 222,422 | 216,956 | 11,101 | 11,101 |
| Strict SNP QC – Prune SNPs on 0.5 max LD | 216,956 | 141,489 | 11,101 | 11,101 |
| Sample QC – Remove bad QC samples | 141,489 | 141,489 | 11,101 | 11,099 |
| Sample QC – Remove mismatch sex pheno samples | 141,489 | 141,489 | 11,099 | 11,061 |
| Sample QC – Remove bad batch samples | 141,489 | 141,489 | 11,061 | 10,979 |
| Sample QC – Remove samples as heterozygosity outliers (+/- 2SD) | 141,489 | 141,489 | 10,979 | 10,232 |

Note: The steps coloured in blue are the QC steps to generate a strict quality control SNP list, which only used to process bad samples exclusion and to calculate PCs. Chr = Chromosome, HWE = Hardy-Weinberg Equilibrium, MAF= Minor Allele Frequency, PC = Principal Component, Pheno = Phenotype, QC = Quality Control, Standard Deviation= SD, SNP = Single Nucleotide Polymorphism

**1.1 Relatedness checking**

From the samples that passed the QC (n = 10,232), a relatedness check was performed (using Identity by descent (IBD) information from plink --genome function). As we can see in Table S4, kinship and relationship of the individuals was checked from the Family ID obtained from the "ABCD ACS Post Stratification Weights" dataset. As shown in Figure 1, the distribution of pi-hat value (Proportion IBD, i.e. P(IBD=2) + 0.5*P(IBD=1)) is within expectation: MZ ~ 1, DZ ~ 0.5 and siblings ~ 0.5. Hence, we did not exclude any samples from this step.

#### **Table S3** Relatedness checking. The number of singletons, siblings, twins and triplets in the sample that passed the QC are reported.

| Relatedness | N |
| --- | --- |
| Singletons | 6,738 |
| Siblings | 1,548 |
| Twins | 1,917 |
| Triplets | 29 |

#### **Figure S1** Violin plot describing $\hat{\pi}$ (pi-hat) value distribution among related individuals.

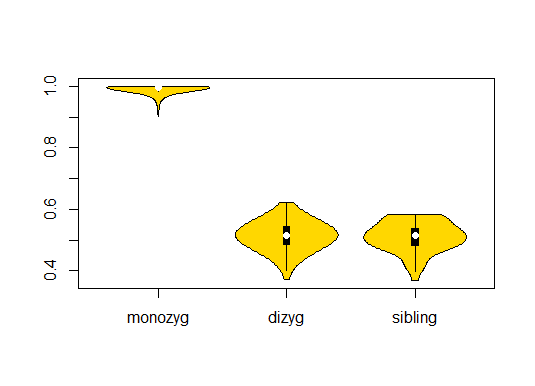

The X axis represents the types of relatedness, categorizing into monozygotic twins (MZ), dizygotic twins (DZ) and siblings. The Y axis represents $\hat{\pi}$ value: (Proportion IBD, i.e. P(IBD=2) + 0.5*P(IBD=1)).

**1.2. Principal Component Analysis**

A Principal Component Analysis (PCA) was performed on a pruned genetic dataset comprising 141,489 LD-independent SNPs, and 20 Principal Components (PCs) were extracted using the plink --pca function.

First of all, in order to gain insights into the distribution of various populations (AFR = Africans, AMR = Admixed Americans, EUR = Europeans, and EAS = East Asians) within the 1000 Genomes Phase 3 Project^10^, PCA plots were conducted. As shown in Figure S2, the area in which the ancestry of each cohort is defined (EUR and AMR) was circled. This area was determined by calculating the mean +/- 3SD of the first two PCs.

Finally, in order to define only those individuals with European ancestry the ABCD and 1000 genomes datasets were merged. As observed in Figures S4 and S7, those individuals within 1000 genomes EUR area were considered to have European ancestry. The number of individuals and % of the total sample are shown in Table S5.

#### **Figure S2** PCA plot of the first 2 PCs of 1000 Genome Phase 3 samples calculated by PLINK 1.9.

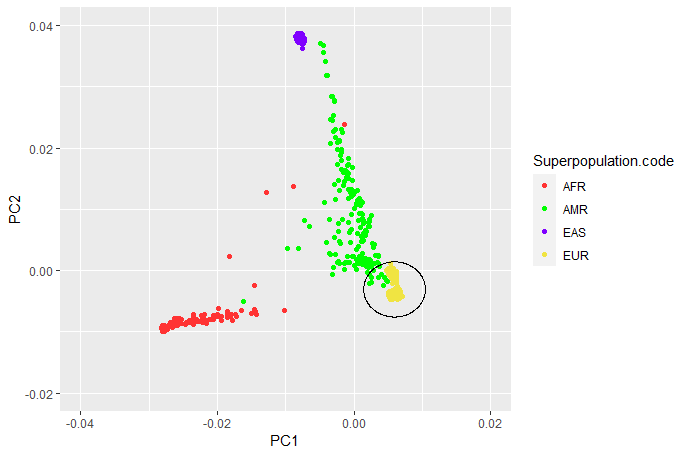

The X and Y axes represent PC1 and PC2 values, respectively. Each population is highlighted in different colours: AFR (Africans, in red, n = 246), AMR (Admixed Americans, in green, n = 181), EAS (East Asians, in purple, n = 286), and EUR (Europeans, in yellow, n = 379). The circled area presents samples defined with European ancestry (mean +/- 3SD for first 2 PCs).

**Figure S3** PCA plot of the first 2 PCs of ABCD samples.

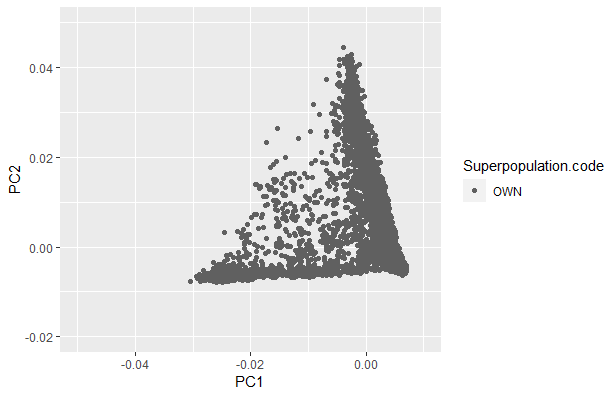

The X and Y axes represent PC1 and PC2 values, respectively. “OWN” cohort is represented (ABCD sample, in grey, n = 10,232). PC = Principal Component

#### **Figure S4** First 2 PCs’ PCA plot of the merged ABCD and 1000 Genomes Phase 3 datasets.

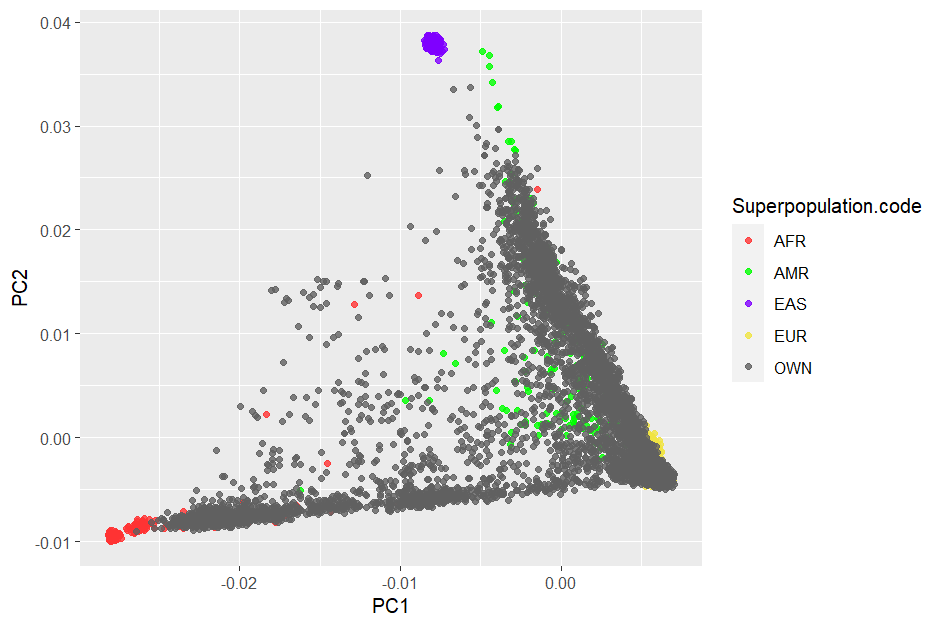

The X and Y axes represent PC1 and PC2 values, respectively. Each population is coloured, and the OWN cohort is represented (ABCD sample, in grey, n = 10,232)

#### **Table S4** Distribution of the ABCD sample per ancestry (number of individuals and % from the total sample)

| Sample Ancestry | | |
| --- | --- | --- |
|  | N | % |
| Total | 10,232 | 100 |
| EUR | 5,656 | 55.28 |
| AMR | 8,352 | 81.63 |
| AFR | 1,195 | 11.68 |

**2. Imputation and Post-imputation Quality Control**

Genotypes where phased (Eagle v.2.4) and imputed to the TOPMed (Version R2 on GRCh38; <https://imputation.biodatacatalyst.nhlbi.nih.gov>) reference panel using MiniMac 4 via the TOPMed Imputation Server,^11^ finally yielding 292,136,462 SNPs. Subsequently, the resulting VCF files were converted into dosage PLINK files and then to hard-call genotypes format.

A post-imputation SNP QC was performed using PLINK 1.9^9^ according to the following criteria: (1) removed SNPs with a MAF < 1%, (2) Imputation Quality Information (INFO) < 0.9, (3) ambiguous and multiallelic SNPs, (4) insertions and deletions variants, and finally (5) SNPs with HWE p-value < 1e-6. The detail of SNP QC steps is shown in Table S6. In the end, there are 10,232 subjects and 5,909,115 SNPs included in the imputed ABCD data.

#### **Table S5** Post-imputation QC describing the SNPs and subjects excluded in all of the steps.

| **SNP QC Step** | **SNP start** | **SNP end** | **Subjects start** | **Subjects end** |
| --- | --- | --- | --- | --- |
| Remove SNPs with MAF < 1% | 292,136,462 | 11,635,718 | 10,232 | 10,232 |
| Remove SNPs with INFO < 0.9 | 11,635,718 | 9,999,328 | 10,232 | 10,232 |
| Remove ambiguous SNPs | 9,999,328 | 8,551,596 | 10,232 | 10,232 |
| Remove multiallelic SNPs | 8,551,596 | 8,224,505 | 10,232 | 10,232 |
| Remove insertions and deletions | 8,224,505 | 7,659,218 | 10,232 | 10,232 |
| Remove SNPs with HWE 1e-6 | 7,659,218 | 5,909,115 | 10,232 | 10,232 |

HWE = Hardy-Weinberg Equilibrium, MAF =Minor Allele Frequency, QC = Quality Control, SNP = Single Nucleotide Polymorphism

**3. PRS calculation**

For target dataset, there were 10,232 subjects and 5,909,115 imputed SNPs included in ABCD data. PRS scores were based on associated alleles and effect sizes reported in the GWAS (Genome-Wide Association Study) summary statistics from the SCZ PGC freeze 3 European subsample^12^.

PRS were generated using the PRS-CS (Polygenic Risk Score-Continuous Shrinkage) software package approach.^13^ For this process, we used the PRS-CS “auto” function, which employs a fully Bayesian approach. This approach automatically learns the global shrinkage parameter(ϕ) from the available data. Unlike other methods, it does not prune SNPs based on a specific p-value threshold or perform clumping for independent SNPs. Instead, it assumes a general distribution of effect sizes across the genome and considers for LD between SNPs using an external LD reference panel (in our case, the 1000 Genomes Phase 3 European samples panel). The SNPs in the base data (n= 7,659,767) underwent QC measures, resulting in 742,011 variants used to calculate PRS. The QC steps involved eliminating multiallelic SNPs, SNPs with bad imputation quality (INFO score below 0.9), ambiguous SNPs, and insertions and deletions variants. Extraction common SNPs from target (ABCD samples), base (GWAS summary statistics) and reference (1000 Genomes Phase 3)^10^ datasets, and subsequently excluding SNPs located in 20 complex-LD regions.^14^

Further details regarding the steps of PRS generation can be found in Table S7.

#### **Table S6** Base Data QC pre-PRS calculation using PRS-cs-auto.

| Step | Base Data |
| --- | --- |
|  | SCZ PGC3 (EUR) |
| Initial SNPs | 7,659,767 |
| SNP QC -- Remove multiallelic SNPs | 7,651,642 |
| SNP QC -- Remove SNPs with INFO <= 0.9 | 6,401,110 |
| SNP QC - Remove ambiguous SNPs | 5,428,447 |
| SNP QC -- Remove insertions and deletions | 5,428,447 |
| Common SNPs between Target, Base and External LD Reference Panel + Remove long LD regions from Target Data | 742,011 |

INFO= Imputation Quality Information, QC = Quality Control,

SCZ = Schizophrenia, SNP = Single Nucleotide Polymorphism
