## Supplementary material for "Sex Differences in the Independent and Combined Effects of Genomic and Exposomic Risks for Schizophrenia on Distressing Psychotic Experiences: Insights from the ABCD Study": Online Resource 2

**Journal:** Archives of Women’s Mental Health

Thanavadee Prachason^1^, Angelo Arias-Magnasco^2^, Bochao Danae Lin^2^, Jim van Os^2-4^, Bart P.F. Rutten^2^, Lotta-Katrin Pries^2^* and Sinan Guloksuz^2,5^*^#^

*^1^Department of Psychiatry, Faculty of Medicine Ramathibodi Hospital, Mahidol University, Bangkok, Thailand.*

*^2^Department of Psychiatry and Neuropsychology, School for Mental Health and Neuroscience, Maastricht University Medical Centre, Maastricht, The Netherlands.*

*^3^Department of Psychiatry, UMC Utrecht Brain Centre, University Medical Centre Utrecht, Utrecht University, Utrecht, The Netherlands.*

*^4^Department of Psychosis Studies, Institute of Psychiatry, Psychology & Neuroscience, King's College London, London, UK.*

*^5^Department of Psychiatry, Yale University School of Medicine, New Haven, Connecticut.*

* Shared last author

**^#^ Corresponding author:** Sinan Guloksuz, Department of Psychiatry and Neuropsychology, School for Mental Health and Neuroscience, Maastricht University Medical Center, P.O. Box 616 6200 MD Maastricht, The Netherlands; tel: 31-433-88-4071, fax: 31433-88-4122,

### **Table S1** Sex differences in the main associations of PRS-SCZ75 with distressing PEs at 3-year follow-up in an unrelated sample

| Distressing PEs | Model 1 | | | | | Model 2 | | | | |
| --- | --- | --- | --- | --- | --- | --- | --- | --- | --- | --- |
|  | Males (N = 2,232) | | Females (N = 1,958) | | Gender difference | Males (N = 2,139) | | Females (N = 1,886) | | Gender difference |
|  | OR  [95% CI] | *P* | OR  [95% CI] | *P* | *P* | OR  [95% CI] | *P* | OR  [95% CI] | *P* | *P* |
| Past-month | 0.93  [0.65 to 1.33] | .695 | 1.23  [0.93 to 1.63] | .149 | .292 | 0.91  [0.63 to 1.32] | .628 | 1.23  [0.92 to 1.64] | .162 | .260 |
| Lifetime  (≥ 1 wave) | 0.98  [0.80 to 1.21] | .844 | 1.42  [1.15 to 1.75] | **.001** | **<.001** | 0.98  [0.79 to 1.21] | .863 | 1.41  [1.13 to 1.75] | **.002** | **<.001** |
| Repeating  ≥ 2 waves | 1.13  [0.85 to 1.51] | .388 | 1.42  [1.09 to 1.85] | **.009** | .247 | 1.12  [0.83 to 1.50] | .457 | 1.40  [1.07 to 1.84] | **.014** | .226 |
| Repeating  ≥ 3 waves | 1.18  [0.76 to 1.84] | .459 | 1.14  [0.77 to 1.70] | .516 | .929 | 1.20  [0.77 to 1.89] | .416 | 1.11  [0.73 to 1.67] | .631 | .822 |
| Persisting  (all 4 waves) | 1.48  [0.66 to 3.33] | .339 | 1.01  [0.53 to 1.91] | .983 | .393 | 1.38  [0.61 to 3.13] | .444 | 0.96  [0.49 to 1.87] | .899 | .431 |

Model 1: Adjusted for age; Model 2: Adjusted for age, family income, and parental education. *p*-values <.05 were bolded. PEs, psychotic experiences; OR, odds ratio; CI, confidence interval.

### **Table S2** Sex differences in the main associations of ES-SCZ75 at 2-year follow-up with distressing PEs at 3-year follow-up in an unrelated sample

| Distressing PEs | Model 1 | | | | | Model 2 | | | | |
| --- | --- | --- | --- | --- | --- | --- | --- | --- | --- | --- |
|  | Males (N = 2,232) | | Females (N = 1,958) | | Gender difference | Males (N = 2,139) | | Females (N = 1,886) | | Gender difference |
|  | OR  [95% CI] | *P* | OR  [95% CI] | *P* | *P* | OR  [95% CI] | *P* | OR  [95% CI] | *P* | *P* |
| ^a^ Past-month | 1.56  [1.14 to 2.15] | **.006** | 1.51  [1.15 to 1.98] | **.003** | .841 | 1.47  [1.05 to 2.05] | **.025** | 1.42  [1.06 to 1.89] | **.017** | .870 |
| Lifetime  (≥ 1 wave) | 2.15  [1.78 to 2.59] | **<.001** | 2.05  [1.67 to 2.50] | **<.001** | .658 | 2.08  [1.71 to 2.53] | **<.001** | 1.88  [1.53 to 2.32] | **<.001** | .410 |
| Repeating  ≥ 2 waves | 2.93  [2.27 to 3.79] | **<.001** | 2.21  [1.73 to 2.82] | **<.001** | .062 | 2.79  [2.13 to 3.65] | **<.001** | 2.03  [1.57 to 2.61] | **<.001** | .050 |
| Repeating  ≥ 3 waves | 3.81  [2.53 to 5.73] | **<.001** | 2.91  [2.04 to 4.16] | **<.001** | .349 | 3.24  [2.12 to 4.94] | **<.001** | 2.64  [1.82 to 3.82] | **<.001** | .501 |
| Persisting  (all 4 waves) | 2.43  [1.15 to 5.15] | **.020** | 3.63  [2.08 to 6.33] | **<.001** | .356 | 2.04  [0.94 to 4.43] | .072 | 3.16  [1.77 to 5.64] | **<.001** | .328 |

Model 1: Adjusted for age; Model 2: Adjusted for age, family income, and parental education

*p*-values <.05 were bolded. PEs, psychotic experiences; OR, odds ratio; CI, confidence interval

^a^ Additionally adjusted for distressing PEs up to 2-year-follow-up

### **Table S3** Joint associations of PRS-SCZ75 and ES-SCZ75 at 2-year follow-up with distressing PEs at 3-year follow-up in unrelated female adolescents

| Distressing PEs |  | Model 1 (N = 1,958) | | | Model 2 (N = 1,886) | | |
| --- | --- | --- | --- | --- | --- | --- | --- |
|  |  | PRS-SCZ75 = 0  OR (95% CI) | PRS-SCZ75 = 1  OR (95% CI) | RERI  (95% CI) | PRS-SCZ75 = 0  OR (95% CI) | PRS-SCZ75 = 1  OR (95% CI) | RERI  (95% CI) |
| ^a^ Past-month (3-year follow-up) | ES-SCZ75 = 0 | 1.0 | 1.02 (0.70 to 1.48)  p = .931 | 0.33  (-0.56 to 1.23)  p = .467 | 1.0 | 1.08 (0.74 to 1.59)  p = .682 | 0.16  (-0.74 to 1.05)  p = .729 |
|  | ES-SCZ75 = 1 | 1.43 (1.04 to 1.98)  **p = .029** | 1.78 (1.12 to 2.83)  **p = .014** |  | 1.39 (0.99 to 1.94)  p = .055 | 1.63 (1.00 to 2.66)  **p = .049** |  |
| Lifetime  (≥ 1 wave) | ES-SCZ75 = 0 | 1.0 | 1.42 (1.10 to 1.83)  **p = .007** | 0.63  (-0.53 to 1.79)  p = .286 | 1.0 | 1.41 (1.08 to 1.82)  **p = .011** | 0.52  (-0.59 to 1.62)  p = .358 |
|  | ES-SCZ75 = 1 | 2.02 (1.60 to 2.55)  **p < .001** | 3.07 (2.13 to 4.43)  **p < .001** |  | 1.86 (1.46 to 2.37)  **p < .001** | 2.78 (1.90 to 4.08)  **p < .001** |  |
| Repeating  (≥ 2 waves) | ES-SCZ75 = 0 | 1.0 | 1.33 (0.95 to 1.87)  p = .098 | 1.03  (-0.36 to 2.42)  p = .146 | 1.0 | 1.31 (0.93 to 1.86)  p = .123 | 0.94  (-0.40 to 2.27)  p = .169 |
|  | ES-SCZ75 = 1 | 2.08 (1.55 to 2.78)  **p < .001** | 3.44 (2.29 to 5.17)  **p < .001** |  | 1.90 (1.40 to 2.57)  **p < .001** | 3.15 (2.06 to 4.81)  **p < .001** |  |
| Repeating  (≥ 3 waves) | ES-SCZ75 = 0 | 1.0 | 1.06 (0.60 to 1.87)  p = .850 | 0.79  (-1.23 to 2.81)  p = .444 | 1.0 | 1.00 (0.56 to 1.81)  p = 991 | 0.78  (-1.12 to 2.67)  p = .421 |
|  | ES-SCZ75 = 1 | 2.75 (1.80 to 4.19)  **p < .001** | 3.59 (2.03 to 6.36)  **p < .001** |  | 2.46 (1.59 to 3.80)  **p < .001** | 3.24 (1.79 to 5.87)  **p < .001** |  |
| Persisting  (all 4 waves) | ES-SCZ75 = 0 | 1.0 | 0.87 (0.32 to 2.42)  p = .795 | 0.64  (-2.72 to 3.99)  p = .711 | 1.0 | 0.92 (0.33 to 2.57)  p = .877 | 0.18  (-2.86 to 3.22)  p = .908 |
|  | ES-SCZ75 = 1 | 3.41 (1.77 to 6.57)  **p < .001** | 3.92 (1.63 to 9.44)  **p = .002** |  | 3.09 (1.56 to 6.11)  **p = .001** | 3.19 (1.24 to 8.21)  **p = .016** |  |

Model 1: Adjusted for age; Model 2: Adjusted for age, family income, and parental education

*p*-values <.05 were bolded. PEs, psychotic experiences; OR, odds ratio; CI, confidence interval

^a^ Additionally adjusted for distressing PEs up to 2-year-follow-up

### **Table S4** Joint associations of PRS-SCZ75 and ES-SCZ75 at 2-year follow-up with distressing PEs at 3-year follow-up in unrelated male adolescents

| Distressing PEs |  | Model 1 (N = 2,232) | | | Model 2 (N = 2,139) | | |
| --- | --- | --- | --- | --- | --- | --- | --- |
|  |  | PRS-SCZ75 = 0  OR (95% CI) | PRS-SCZ75 = 1  OR (95% CI) | RERI  (95% CI) | PRS-SCZ75 = 0  OR (95% CI) | PRS-SCZ75 = 1  OR (95% CI) | RERI  (95% CI) |
| ^a^ Past-month (3-year follow-up) | ES-SCZ75 = 0 | 1.0 | 0.71 (0.41 to 1.22)  p = .212 | 0.47  (-0.45 to 1.38)  p = .321 | 1.0 | 0.72 (0.42 to 1.25)  p = .245 | 0.38  (-0.50 to 1.27)  p = .397 |
|  | ES-SCZ75 = 1 | 1.48 (1.02 to 2.15)  p = .037 | 1.65 (0.99 to 2.76)  p = .055 |  | 1.40 (0.95 to 2.05)  p = .088 | 1.50 (0.88 to 2.56)  p = .136 |  |
| Lifetime  (≥ 1 wave) | ES-SCZ75 = 0 | 1.0 | 0.87 (0.66 to 1.14)  p = .298 | 0.48  (-0.32 to 1.28)  p = .236 | 1.0 | 0.87 (0.66 to 1.15)  p = .316 | 0.49  (-0.31 to 1.29)  p = .230 |
|  | ES-SCZ75 = 1 | 2.00 (1.61 to 2.48)  **p < .001** | 2.35 (1.70 to 3.24)  **p < .001** |  | 1.93 (1.54 to 2.42)  **p < .001** | 2.29 (1.64 to 3.19)  **p < .001** |  |
| Repeating  (≥ 2 waves) | ES-SCZ75 = 0 | 1.0 | 0.98 (0.64 to 1.50)  p = .912 | 0.85  (-0.58 to 2.27)  p = .245 | 1.0 | 0.95 (0.61 to 1.48)  p = .832 | 0.83  (-0.56 to 2.22)  p = .244 |
|  | ES-SCZ75 = 1 | 2.79 (2.07 to 3.78)  **p < .001** | 3.62 (2.41 to 5.42)  **p < .001** |  | 2.65 (1.93 to 3.63)  **p < .001** | 3.43 (2.26 to 5.21)  **p < .001** |  |
| Repeating  (≥ 3 waves) | ES-SCZ75 = 0 | 1.0 | 1.08 (0.52 to 2.24)  p = .838 | 0.83  (-1.80 to 3.46)  p = .536 | 1.0 | 1.05 (0.51 to 2.19)  p = 888 | 0.91  (-1.44 to 3.26)  p = .448 |
|  | ES-SCZ75 = 1 | 3.78 (2.34 to 6.10)  **p < .001** | 4.69 (2.55 to 8.63)  **p < .001** |  | 3.17 (1.93 to 5.21)  **p < .001** | 4.13 (2.22 to 7.69)  **p < .001** |  |
| Persisting  (all 4 waves) | ES-SCZ75 = 0 | 1.0 | 1.98 (0.63 to 6.19)  p = .239 | -0.49  (-5.19 to 4.21)  p = .838 | 1.0 | 1.91 (0.61 to 6.00)  p = .266 | -0.83  (-4.92 to 3.26)  p = .690 |
|  | ES-SCZ75 = 1 | 3.25 (1.28 to 8.29)  **p = .014** | 3.74 (1.09 to 12.9)  **p = .036** |  | 2.84 (1.09 to 7.42)  **p = .033** | 2.92 (0.81 to 10.5)  p = .100 |  |

Model 1: Adjusted for age; Model 2: Adjusted for age, family income, and parental education

*p*-values <.05 were bolded. PEs, psychotic experiences; OR, odds ratio; CI, confidence interval

^a^ Additionally adjusted for distressing PEs up to 2-year-follow-up

### **Table S5** Sex differences in the additive interaction between PRS-SCZ75 and ES-SCZ75 at 2-year follow-up on distressing PEs at 3-year follow-up in an unrelated sample

| Distressing PEs | Model 1 | | | | | Model 2 | | | | |
| --- | --- | --- | --- | --- | --- | --- | --- | --- | --- | --- |
|  | Males (N = 2,232) | | Females (N = 1,958) | | Gender difference | Males (N = 2,139) | | Females (N = 1,886) | | Gender difference |
|  | RERI  [95% CI] | *P* | RERI  [95% CI] | *P* | *P* | RERI  [95% CI] | *P* | RERI  [95% CI] | *P* | *P* |
| ^a^ Past-month | 0.47  [-0.45 to 1.38] | .321 | 0.33  [-0.56 to 1.23] | .467 | .803 | 0.38  [-0.50 to 1.27] | .397 | 0.16  [-0.74 to 1.05] | .729 | .661 |
| Lifetime  (≥ 1 wave) | 0.48  [-0.32 to 1.28] | .236 | 0.63  [-0.53 to 1.79] | .286 | .840 | 0.49  [-0.31 to 1.29] | .230 | 0.52  [-0.58 to 1.62] | .358 | .972 |
| Repeating  ≥ 2 waves | 0.85  [-0.58 to 2.27] | .245 | 1.03  [-0.36 to 2.42] | .146 | .830 | 0.83  [-0.56 to 2.22] | .244 | 0.94  [-0.40 to 2.27] | .169 | .906 |
| Repeating  ≥ 3 waves | 0.83  [-1.80 to 3.46] | .536 | 0.79  [-1.23 to 2.81] | .444 | .983 | 0.91  [-1.44 to 3.26] | .448 | 0.78  [-1.12to 2.67] | .421 | .938 |
| Persisting  (all 4 waves) | -0.49  [-5.19 to 4.21] | 0.838 | 0.64  [-2.72 to 3.99] | .711 | .669 | -0.83  [-4.92 to 3.26] | .690 | 0.18  [-2.86 to 3.22] | .908 | .641 |

Model 1: Adjusted for age; Model 2: Adjusted for age, family income, and parental education

*p*-values <.05 were bolded. PEs, psychotic experiences; OR, odds ratio; CI, confidence interval

^a^ Additionally adjusted for distressing PEs up to 2-year-follow-up
